# A longitudinal study of the impact of human mobility on the incidence of COVID-19 in India

**DOI:** 10.1101/2020.12.21.20248523

**Authors:** Sarbeswar Praharaj, Hoon Han

## Abstract

Human mobility plays a crucial role in determining how fast and where infectious diseases can spread. This study aims to investigate visit to which category of places among grocery, retail, parks, workplaces, residential, and transit stations is more associated with the incidence of COVID-19 in India. A longitudinal analysis of generalized estimating equation (GEE) with a Poisson log-linear model is employed to analyze the daily mobility rate and reported new cases of COVID-19 between March 14 and September 11, 2020. This study finds that mobility to places of grocery (food and vegetable markets, drug stores etc.) and retail (restaurants, cafes, shopping centres etc.) is significantly associated (at p<0.01) with the incidence of COVID-19. In contrast, visits to parks, transit stations and mobility within residential neighbourhoods are not statistically significant (p>0.05) in changing COVID-19 cases over time. These findings highlight that instead of blanket lockdown restrictions, authorities should adopt a place-based approach focusing on vulnerable hotspot locations to contain the COVID-19 and any future infectious disease.

## Introduction

The globalized world, in which movement of people has increased at an unprecedented scale is susceptible to the spread of infectious disease on a global and local scale (Tatem et al., 2006). With rapid advancements in transportation systems, our societies have increased reach, speed of travel and human beings are moving in great volumes like never before. The global spread of the new coronavirus SARS-CoV-2-pathogen, also known as COVID-19 (Chen et al., 2020; Lai et al., 2020; Singhal, 2020; Sohrabi et al., 2020; D. Wang et al., 2020; L. Wang et al., 2020) that has led to over 70 million infections worldwide as of Mid-December 2020 (WHO, 2020a) is indicative of how mobility can turn into a global health problem in an increasingly globalized world. A popular model adopted across the global regions to curb the rise of infections is enforcing social distancing measures (Cato et al., 2020; Lasry et al., 2020; Lunn et al., 2020; WHO, 2020b; Zhu et al., 2020), including bans on non-essential travel, stay-at-home orders, school closures, and temporary shutdown of offices and businesses. These country-based control and mitigation measures somewhat determined the course of the COVID-19 pandemic (Anderson et al., 2020).

India is the second-worst hit country by the COVID-19 health crisis as of December 19, 2020, touching 10 million cases and more than 1.4 million people succumbing to the virulent disease (WHO, 2020a). India’s first COVID-19 case was reported on January 30, 2020, in the state of Kerala, where the patient had travelled from Wuhan in China, the original epicentre of the epidemic (Rafiq et al., 2020). At the early stages of the pandemic, the Government of India announced an unprecedented nationwide lockdown on March 24, 2020, banning non-essential travel within the country and cancelling international flights to prevent the spread of COVID-19. The lockdown on 1.38 billion Indians was aimed to enforce strict social distancing and was implemented in four phases over nine weeks (March 25 – May 31). The first three weeks of the lockdown were the most severe with nearly all services and factories suspended across the country (Praharaj and Vaidya, 2020). During the second phase of the intervention (April 15 – May 3) lockdown areas were classified into three zones – red (indicating infection hotspots), orange (indicating some infection) and green (no cases of COVID-19 (Praharaj et al., 2020). Some relaxations were announced for orange and green zones in this phase, allowing agricultural businesses, banks, and government centres to reopen. The third (May 4 – 17) and fourth (May 18 – 31) phase of the lockdown was used to further open up activities in the orange zone and to restore normalcy in the green zone. While the number of cases continued to increase in several parts of India since the lockdown was lifted on May 31, 2020, we are yet to understand what impact the mobility restrictions, lockdowns and subsequent reopenings may have had on the evolving scenario of virus spread and the COVID-19 health crisis.

We hypothesize that by studying human mobility data, it is possible to explain the nature of change in individuals’ movements due to social distancing interventions and how that affected the progression of cases over time. Numerous initiatives are tracking mobility changes during the pandemic, such as Google Community Mobility Reports (Google, 2020) and Apple Mobility Trends Reports. These initiatives use anonymized, aggregated data to chart movement trends over time by geography, as well as by place categories, including grocery, retail, workplaces, parks, transit centres and places of residence. These open datasets offer novel opportunities to study the relationship between human mobility patterns and the transmission of COVID-19. The usability and value of mobility data in epidemiology studies are well-established in the literature (Wesolowski et al., 2015, 2012; Wu et al., 2020). Researchers have also used human mobility data as an indicator of social distancing, which is known to have a significant impact on breaking the chain of human-to-human transmission of the virus (Badr et al., 2020; Lasry et al., 2020; WHO, 2020c).

In an attempt to explain the COVID-19 outbreak in the U.S., Paez (Paez, 2020) used the mobility data from GCMR to quantify the regression effects of categories of places, and found that workplace-related trips had the most effect on increased caseload. The same GCMR data was used by Lasry et al. (Lasry et al., 2020) to examine the temporal correlations between public-policy induced change in community mobility and growth of infections to suggest there was an association at least in the Four U.S. Metropolitan Areas that they studied. Badr et al. (Badr et al., 2020) developed a mobility ratio using similar cell phone data to find that falling mobility trends were strongly correlated with decreased COVID-19 case growth rates over time for the most affected counties in the U.S. between January 1 and April 20. Using Apple mobility data Hadjidemetriou et al. (Hadjidemetriou et al., 2020) investigated the impact of the U.K. government’s interventions on human mobility and its subsequent impact on severe COVID-19 outcomes. Mobile location data was used by researchers in China too (Fang et al., 2020; Kraemer et al., 2020), where they found that the reduction in non-essential travel imposed on the epicentre of the epidemic in Wuhan dramatically changed the trajectory of the pandemic.

This study investigates the potential of GCMR data to assess the impact of human mobility on the incidence of COVID-19 in India. We used Google data because Google Map is used extensively in India, and the dataset is potentially more representative of actual mobility patterns. Our study examines one central research question: Is there a relationship between daily mobility patterns and the new incidence of COVID-19? We subsequently explore whether there is a difference in the ways travel to various categories of places like parks, grocery and food markets, and workplaces influence the coronavirus transmission. We were keen to learn, if there is a statistically significant difference in the effects of the different mobility categories, then which are the locations/places that are more prone to spread the infection.

## Data and Methods

### Variables and data sources

We used two datasets for the longitudinal analysis in this study to answer the research questions. We obtained the first set of data – the daily state-level counts of COVID-19 cases (dependent variable) from [COVID-19 India Tracker](https://www.covid19india.org/) (COVID-19 India Org Data Operations Group, 2020), a crowd-sourced database for real-time COVID-19 statistics and patient tracing in India. The dataset is made available through an open (CC-BY-4.0) license at https://api.covid19india.org. This is the most authoritative source available for COVID-19 related information in India. It combines both the daily data from the Indian Ministry of Health and Family Welfare and data published through state health department bulletins from all the 36 states and union territories in India.

Daily mobility indicator data is the second set of data used in the analysis, obtained from Google [Community Mobility Reports](https://www.google.com/covid19/mobility/) (GCMR). GCMR (Google, 2020)provides aggregated and de-identifiable information of Google Map users on their visits to different categories of places based on their location history. Google constructed the data based on the frequency and length of visits to places, and reports percentage change from a baseline level, corresponding to the median value of mobility of identical days of the week during January 3 – February 6, 2020 (Aktay et al., 2020). Of the six categories of places, retail and recreation data provides information on visits to places like restaurants, cafes, shopping centres, libraries and movie theatres. Grocery and pharmacy data indicates mobility trends on visits to grocery markets, food warehouses, farmers markets, drug stores and pharmacies. Parks data includes travel to and from places like national parks, public beaches, plazas, public gardens and marinas. Transit station data provides information on mobility trends on visits to bus and train stations. Workplace data contains information on mobility trends for the location of work, and residential data has trends for movements within neighbourhoods of residence.

Combining the two datasets determined the daily sample for 36 states and union territories in India, spanning six months between March 14 and September 11, 2020. We have made the research data available through the Mendeley Data at http://dx.doi.org/10.17632/wpywf5ny6j.1. The incubation period of COVID-19, which is the time between exposure to the virus and symptom onset, is on average 5– 6 days but can be as long as 14 days as observed by WHO (WHO, 2020d) and the Government of India (Government of India, 2020) based on several scientific publications (Backer et al., 2020; Lauer et al., 2020; Yu et al., 2020). Given this, it is expected that any changes in mobility will have a lagged effect on the discovery of new cases. To counter this, we calculated the lagged moving averages of the mobility indicators. We created both a 7-day lagged indicator and a 14-day lagged indicator to help understand whether a longer incubation time leads to a different prediction and inventory of COVID-19 cases. The lagged indicators are calculated as the mean of the mobility indicator using the values from date-minus-7-days to date-minus-2-days for the 7-day lagged indicator. Furthermore, it is possible that mobility and reports of new cases of COVID-19 are endogenous, if communities adjust their mobility according to increasing incidence rates (Paez, 2020). Hence, in addition to being consistent with a scientifically proven incubation period, the use of lagged indicators also helps to break this potential endogeneity.

### Statistical model

We used a Poisson log-linear model with a generalised estimating equation (GEE) approach, which is a reliable statistical method for longitudinal data analysis where the broad scientific objective is to describe an outcome (Ballinger, 2004), in this case, the incidence of COVID-19. In longitudinal data, repeated observations for a subject tend to be correlated. GEEs use the generalised linear model to estimate more efficient and unbiased regression parameters relative to ordinary least squares regression because the model allows the development of a working correlation matrix (Zeger et al., 1988). This accounts for the within-subject correlation of responses on dependent variables of a variety of distribution types – normal, binomial and Poisson. Initially developed by Zeger and Liang (Zeger and Liang, 1986), the GEE method has been widely used in medical and life sciences. We define the daily COVID-19 case counts for different regions as the dependent variable. This model assumes that the responses are not normally distributed (Poisson distribution) because they consist of a count of the daily infections by place. We used the longitudinal marginal model to quantify the relationship between the incidence of daily COVID-19 cases and the six mobility types (retail and recreation, grocery and pharmacy, parks, transit stations, workplaces and residential) using the correlation among repeated case counts for 36 Indian regions. The model presents a quasi-likelihood estimate of β coefficient arising from the maximisation of normality-based log-likelihood of COVID-19 cases by geography.

The Poisson log-linear regression model for the expected rate of occurrence of COVID-19 cases in a state or territory can be denoted by:

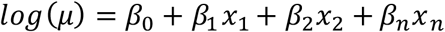

The longitudinal data of COVID-19 cases is daily cases for a particular region. We denote the expected cases, log(μ) and six mobility rates *x*_*n*_ = *x*_1_ + *x*_2_ + *x*_3_ + *x*_4_ + *x*_5_ + *x*_6_.

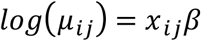

Here, GEE treats the response vector for the daily COVID-19 cases with the mean vector noted by *μ*_*ij*_, which corresponds to *j*^th^ mean. The cases are assumed to be independent across regions but correlated within each region.

## Analysis and results

### COVID-19 outbreak and the containment response in India

Although the COVID-19 lockdown in India was one of the harshest in the world (Daniyal, 2020), the level of implementation and observance by communities across the regions varied. We mapped the progression of cases in different states alongside various policy events in Figure 1. The analysis shows India had a meagre 571 cases on March 24 when the nationwide lockdown started. Before the lockdown was enforced, India’s COVID-19 cases were doubling every three days (Rafiq et al., 2020). At the end of the initial lockdown period on April 18, the growth rate of infections had significantly slowed to doubling every eight days. Our data reveals that by the end of the first two phases of the lockdown on May 3, less than 43,000 total cases were reported across India. As discussed in the introduction, the first two phases of the lockdown were rigorously implemented and, as a result, the authorities were able to manage the virus spread. India experienced a steady rise in cases during the third and fourth phases of the lockdowns, reaching a total of 190,000 cases by the end of May. During these two lockdown phases, mobility restrictions were gradually eased in areas outside the containment zones. The Indian Council for Medical Research (Indian Council of Medical Research (ICMR), 2020) highlighted that at the end of the lockdown, community transmission of the virus had still not been reported in India, due to the pro-active social distancing and mobility restrictions observed during the period.

**Figure 1:**
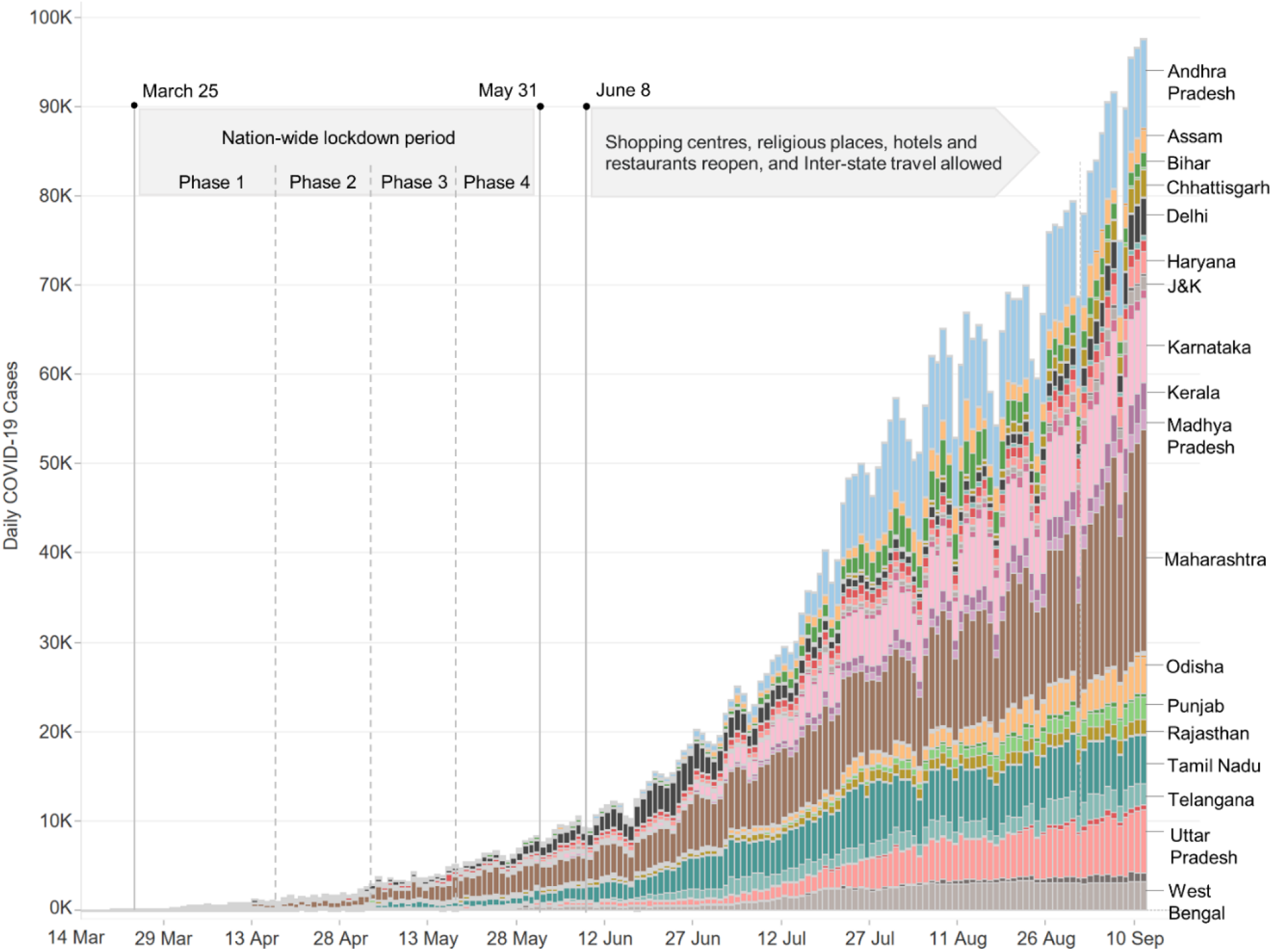
Daily COVID-19 cases by states in India and the timeline of lockdown events

The COVID-19 trajectory in India changed rapidly from mid-June following executive orders to reopen shopping centres, religious places, hotels and restaurants beginning June 8. Restrictions on interstate travel were also withdrawn, which significantly changed mobility dynamics. With the opening up of economic activities and increased movements, there was an alarming increase of COVID-19 infections in new territories like Andhra Pradesh, Tamil Nadu, and Karnataka, as highlighted in Figure 1. India became the country second worst-affected by the COVID-19 pandemic on September 6 when it had 4.2 million total cases. Currently, a large number of daily infections is being reported in densely populated states, including West Bengal, Bihar, and Uttar Pradesh, which were at the forefront of massive migrant crisis during the lockdown. The crisis was driven by a mass exodus of migrant workers and informal labourers who were travelling back to their native states from big cities like Mumbai and Delhi. Thousands of migrant workers, having lost their jobs and facing severe food insecurity (Bhowmick, 2020) due to lockdowns flocked around train and bus stations, and many even walked hundreds of kilometres to return to their native places. Social distancing was not possible for these migrants since they travelled together in large groups, and initially, there was no coordinated response by governments to deal with the migrant crisis. Special trains and buses were operated to transport these stranded workers where intense overcrowding was reported. With India having more than 90,000 new cases a day in early September, the Indian Medical Association revealed that community transmission (WHO, 2020d; Yu et al., 2020) of COVID-19 was in effect in many parts of the country, which means that the pandemic was so widespread that the authorities can no longer identify the chain of virus transmission and that the virus was moving freely in the community.

### Mobility effects by categories of places on the incidence of COVID-19

We developed a GEE model with Poisson log-linear analysis with daily COVID-19 cases as the dependent variable. The analysis was performed through SPSS, and the results of the model are shown in Tables 1 and 2. Table 1 presents the Chi-Square hypothesis test outcomes with 7-day lagged mobility indicators. Table 2 summarises the results for 14-day lagged mobility indicators. We find that mobility related to retail and recreation and grocery and pharmacy are significantly associated (at p<0.01) with the incidence of COVID-19 (see Table 1). The two mobility categories have a positive coefficient (β) of 0.35 and 0.38, respectively, suggesting that an increase in the covariate will result in an increased number of COVID-19 cases over time. The model, therefore, shows that an increase in mobility to places like shopping centres, grocery markets, restaurants, farmers markets and pharmacies results in an increased number of COVID-19 cases in India. We also discover that the coefficients (B) of parks, transit stations and residential related mobility are not statistically significant (p>0.05), which implies that they do not affect changing COVID-19 cases over time. The test results demonstrate that although workplace-related mobility is significantly associated with the dependent variable (p<0.05), the coefficient value for predicting the relationship is negative. We can hence verify that reports of COVID-19 cases due to workplace-related mobility have reduced over time.

**Table 1:**
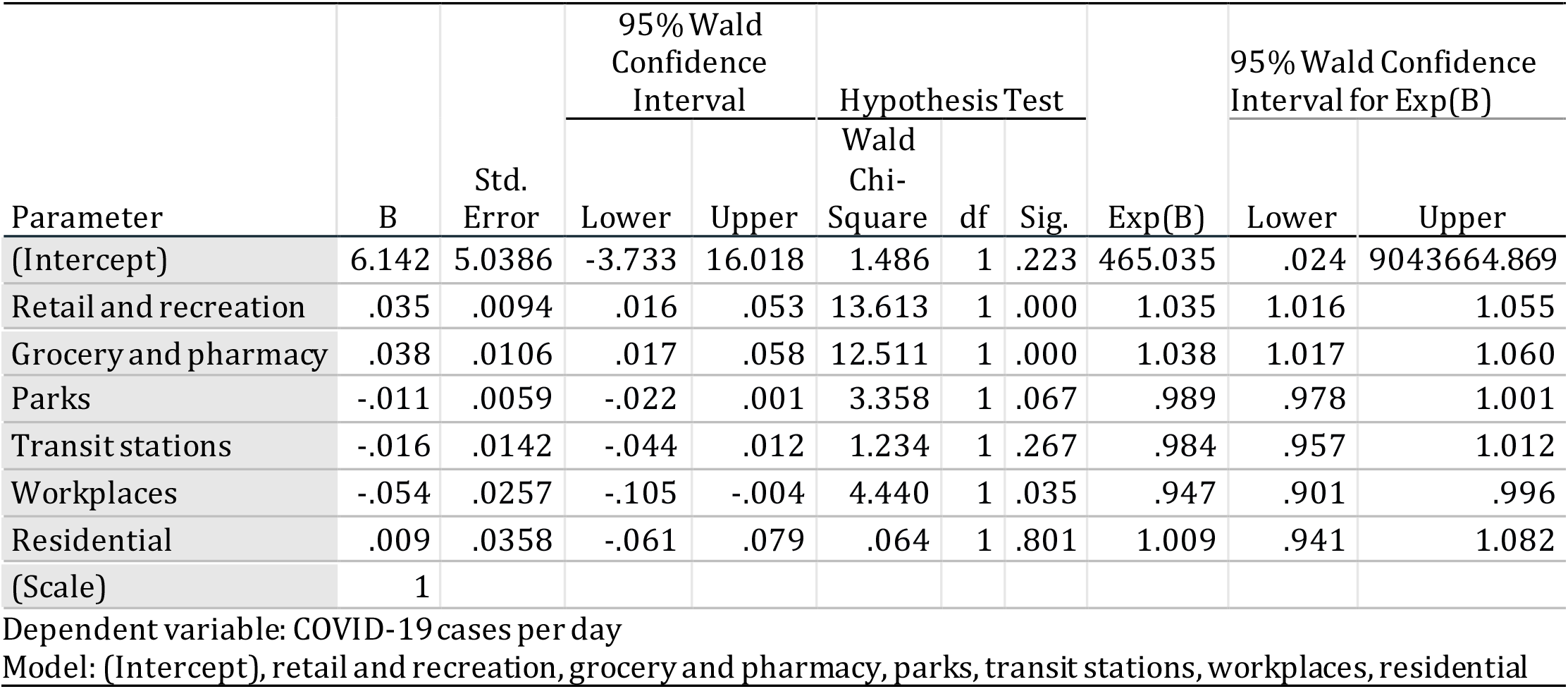
Parameter estimates for 7-day lagged mobility indicators

**Table 2:**
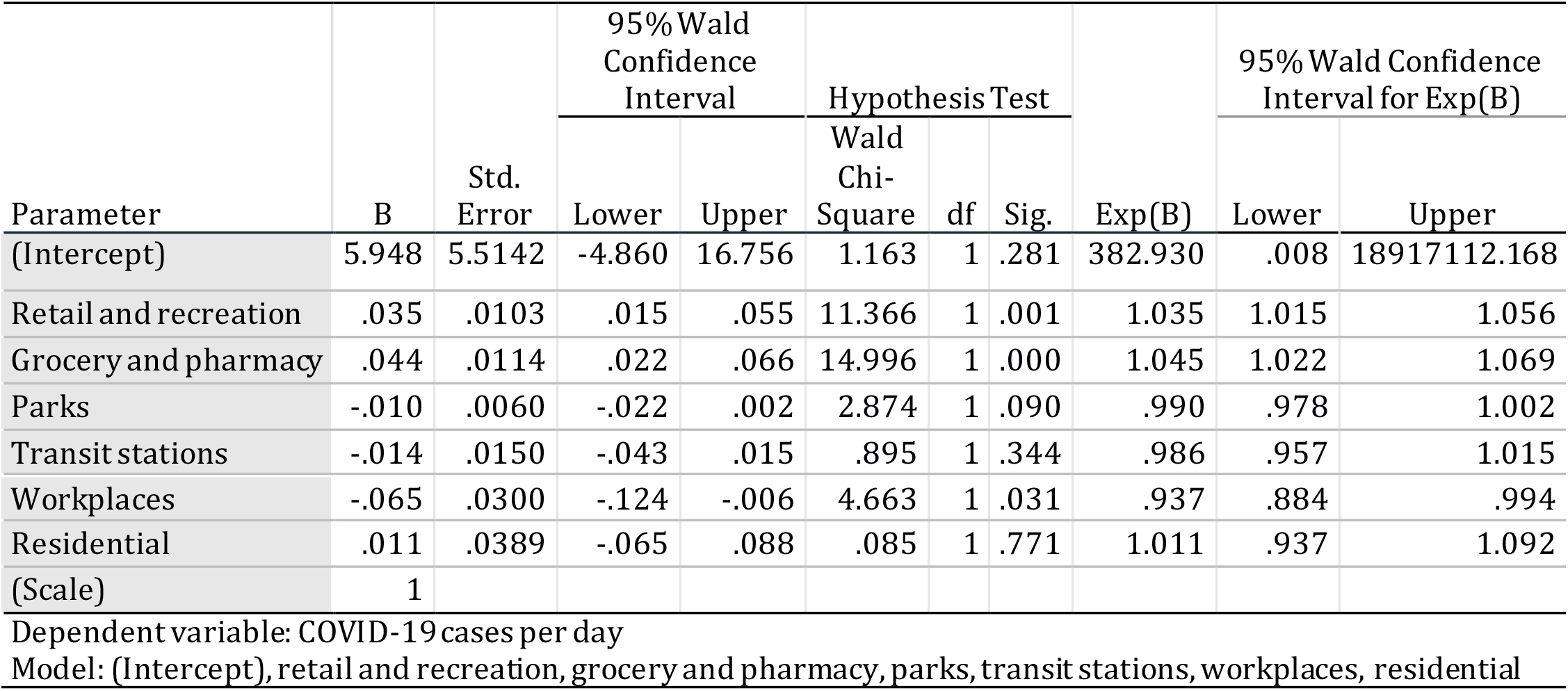
Parameter estimates for 14-day lagged mobility indicators

The GEE Poisson log-linear model results using 14-day lagged mobility indicators are presented in Table 2, showing a very similar outcome to Table 1, which used the 7-day lagged mobility indicators. Thus, we reconfirm the mobility effects discussed in the previous paragraph. In fact Table 2 indicates a significantly stronger coefficient estimate (B=0.044) than Table 1 for mobility related to grocery and pharmacy, re-affirming that visits to grocery markets, food warehouses, farmers markets, speciality food shops, drug stores, pharmacies and similar places increase the chances of coronavirus infection. The use of 14-day lagged mobility indicators also reveals a more substantial negative coefficient value (B=-0.065) for workplace-related travel, highlighting that it poses the lowest risk of transmission of the virus among the six place categories. Detailed SPSS output results are presented in Table 1.1 and 2.1 as supplementary files.

## Discussion

This study presents a novel approach to study the relationship between daily mobility and the incidence of COVID-19 using the GEE method. Our statistical model effectively negates the within-subject correlation of responses on dependent variables to produce unbiased parameter estimates for case count data that was not normally distributed. However, there may be some possible limitations/bias in the data used for analysis, and the results of this study should be interpreted in light of those limitations. The dependent variable consists of daily COVID-19 cases by regions. The availability of testing centres and the number of tests performed across states in India varies significantly (Rafiq et al., 2020; Sarkar et al., 2020). India’s testing rate is slightly above 70,000 per million population as of September 11 (the last date for which data is included in this analysis), which is much lower than other COVID-19 worst-hit countries like USA or Brazil, which have conducted over 400,000 tests per million people (WHO, 2020c). Hence, the actual cases in India, overall and across regions, might be much higher than reported COVID-19 positive cases. Also, depending on the late development or non-availability of testing equipment at the early stages of the pandemic, the accuracy of daily new cases might have been affected. With the mobility indicators, a major limitation was that it includes data for select people who use Google products on a smartphone (Aktay et al., 2020). Therefore, the data might not representatively capture all groups, such as older adults and the low-income population who might not be using smartphones while navigating through different places. The other limitation is to the aggregated number of travels which does not reflect the frequency of travels per person, especially those who travel multiple places and multiple times per day. However, due to the limited accessibility of a large volume of daily travel data in countries such as India, the GPS based open-source mobility statistics provide a robust understanding of COVID-19 cases and mobility patterns at the regional level.

The study establishes that there was a strong relationship between human mobility patterns and the incidence of COVID-19 in India. Importantly, the results highlight that the spread of the coronavirus is closely related to the frequency of short-distance travel for daily needs. The crowdedness of local public places like local grocery and retail shops emerged as the likely key factor that drives human-to-human transmission of coronavirus. The local food and vegetable markets in Indian cities are highly unorganised places, and they lack space for proper social distancing. During the lockdown period, the majority of states restricted the operating hours of these markets, and vendors were only allowed to open for a few hours in the morning. These rules may have had an adverse impact, leading to overcrowding of these market areas due to the influx of a large number of people in a limited time period seeking essential food items. Furthermore, these crowded markets are often located within pockets of poverty, suggesting that the socioeconomic dynamics of localities might have influenced how the virus spread through communities. The findings support previous research (Hamidi et al., 2020; Hawkins et al., 2020; C. Wang et al., 2020), which have established that socioeconomic factors play an important role in COVID-19 prevalence in the USA.

We computed and mapped the mean monthly mobility levels from GCMR data for grocery and pharmacy-related trips to corroborate the model results. A comparison of the mobility levels in April (during lockdown Phase 1) and June (after the lockdown ended on May 31) in Figure 2 highlight that human movements significantly increased in several parts of India since the lockdown was lifted. States like Bihar, Chhattisgarh, Kerala, Uttar Pradesh, West Bengal, Assam were having much higher levels of mobility even during the lockdown in April. Most of these regions were at the forefront of the migrant crisis, where labourers in large numbers returned from other states due to joblessness and hunger. Interestingly these are the regions which show a manifold increase in new COVID-19 cases since the relaxing of lockdown restrictions as shown in Table 3, where we capture the progression of case counts across states over a six-month period (March 14 to September 11). For example, Bihar added over 150,000 cases between July 11 and September 11, whereas it had less than 1,000 total cases till May 11. Similarly, in West Bengal, nearly 200,000 cases occurred in the two-month period between July 11 and September 11, while there were only 5,500 cases on May 31.

**Table 3:** New cases (in thousands) reported in the most-affectedstates over six months

| States | Mar 14 - May 11 | May 11 - July 11 | July 11 - Sep 11 |
| --- | --- | --- | --- |
| Andhra Pradesh | 2.02 | 27.24 | 547.69 |
| Assam | 0.07 | 15.97 | 138.24 |
| Bihar | 0.75 | 15.04 | 155.45 |
| Chhattisgarh | 0.06 | 3.90 | 58.64 |
| Delhi | 7.23 | 110.92 | 209.75 |
| Gujarat | 8.54 | 41.03 | 110.97 |
| Haryana | 0.73 | 20.58 | 88.33 |
| Jharkhand | 0.16 | 3.66 | 59.02 |
| Karnataka | 0.86 | 36.22 | 440.41 |
| Kerala | 0.52 | 7.44 | 102.25 |
| Madhya Pradesh | 3.79 | 17.20 | 83.62 |
| Maharashtra | 23.40 | 246.60 | 1015.68 |
| Odisha | 0.42 | 12.53 | 143.12 |
| Punjab | 1.88 | 7.59 | 74.62 |
| Rajasthan | 3.99 | 23.75 | 99.04 |
| Tamil Nadu | 8.00 | 134.23 | 491.57 |
| Telangana | 1.28 | 33.40 | 152.09 |
| Uttar Pradesh | 3.57 | 35.09 | 299.05 |
| West Bengal | 2.06 | 28.45 | 196.33 |
| India total | 85.86 | 884.30 | 3773.10 |

**Figure 2:**
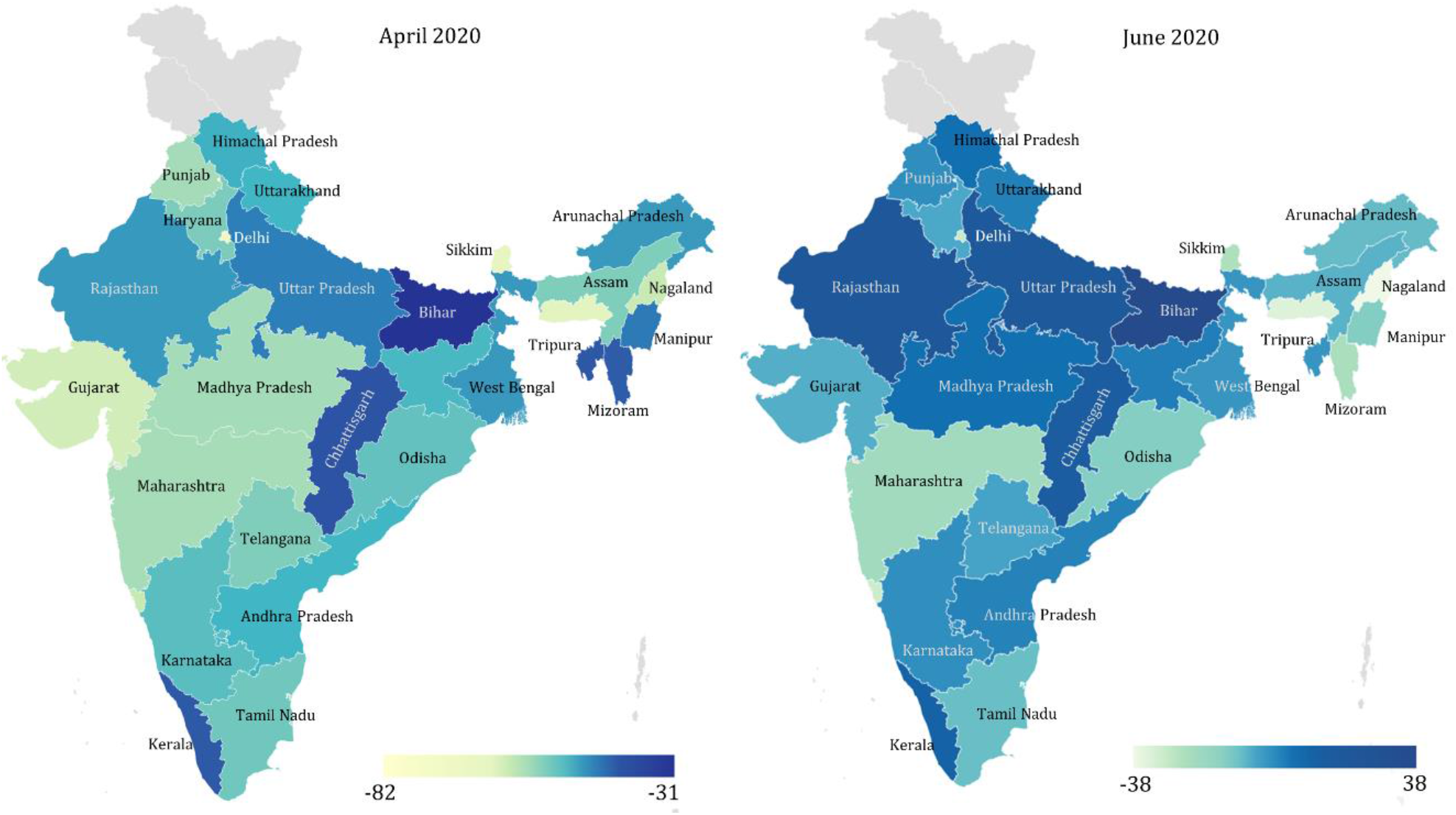
Comparison of mean monthly mobility levels for grocery & pharmacy-related trips

## Conclusion

The findings strongly suggest that better crowd management and social distancing strategies in local markets and shopping districts is critical to check the spread of the COVID-19 and any future infectious disease. The study indicates the need for a more cohesive approach to managing the interstate and city-to-village movements of migrant workers to address the threat of community transmission during such future health crisis. We also found that blanket lockdown restrictions might not be beneficial, as this research provides evidence that economic activities like commuting to workplaces, which have preventive measures such as social distancing and wearing of masks, do not lead to an increased number of cases. Likewise, we find that visiting low-density open spaces (public parks) does not significantly affect virus spread. Therefore, social distancing, which has been established by the previous researchers (Badr et al., 2020; Cato et al., 2020; Lasry et al., 2020; Singhal, 2020; WHO, 2020c) as a critical tool in the battle against COVID-19, must be continued to be rigorously implemented in the evolving context of the pandemic to control any subsequent waves of infections. Overall, the results from this study show that initial control measures and lockdown aimed at human mobility reduction had a decent impact on the occurrence of COVID-19 in India. However, given the limited resources available for enforcing extensive lockdowns in a large country and its adverse effects on the economy, authorities should adopt a place-based approach, focusing on promoting and implementing strict social-distancing in hotspot locations that are most vulnerable.

The result is original and, if confirmed in other case studies, would lay the groundwork for more effective containment of COVID-19 in India and other countries that are still experiencing a health emergency. The findings and lessons from this study shall help design and evolve a better COVID-19 response during the ongoing crisis as well as for planning ahead to minimise the impacts of likely subsequent waves of the virus, or for other pandemics. Further research should examine the mobility effects on the incidence of COVID-19 at the different hierarchy of places within regions, e.g. urban areas, inner cities, and peripheral rural areas using more disaggregated data. Additionally, future work may investigate socio-economic and demographic factors, such as age, poverty rates, income, race, population density, and housing condition to understand which individual measures or combination of them alongside human mobility increase the vulnerability of places to occurrences of COVID-19 or pandemics alike.

## Data Availability

We have made the research data available through the Mendeley Data at http://dx.doi.org/10.17632/wpywf5ny6j.1

## Acknowledgements

The authors would like to acknowledge the open data providers - Google and COVID-19 India, which enabled us to perform the analysis presented in this paper.

## Ethical approval

Not required. Both the dataset used in this study came from a public database. The Google COVID-19 Community Mobility Reports data was collected and used in accordance to the to the Google [Terms of Service](https://policies.google.com/terms?hl=en). Whereas the COVID-19 India dataset was accessed through an open API Creative Commons Attribution 4.0 International License.

